# Evidence of Accumulating Neurophysiologic Dysfunction in Persistent Post-COVID Fatigue

**DOI:** 10.64898/2025.12.19.25342669

**Authors:** Maria Germann, Natalie J. Maffitt, Olivia A. Burton, Amn Ashhad, Anne M.E. Baker, Boubker Zaaimi, Wan-Fai Ng, Demetris S. Soteropoulos, Stuart N. Baker, Mark R. Baker

## Abstract

A major consequence of the COVID-19 pandemic has been the emergence of post-COVID syndrome (PCS), and more specifically, post-COVID fatigue (pCF), with an estimated prevalence of ∼2%. We previously showed that, compared to healthy controls, people with pCF exhibit changes in muscle physiology, cortical circuitry, and autonomic function.

Here we present results from a cohort of people with pCF (N=145), between 12 weeks and 45 months post-infection. We report self-perception of fatigue; objective measures of cortical circuits via transcranial magnetic stimulation and reaction time tasks; peripheral muscle fatigue; and autonomic function such as heart rate variability.

Those with pCF persisting >200 days had significantly more fatigue and showed increased cortical excitability, slower reaction times and increased peripheral muscle fatigue compared to those with <200 days of pCF .

In pCF, if there is no spontaneous recovery, fatigue worsens, and patients continue to accumulate significant neurophysiologic abnormalities.

## Introduction

Since the onset of the COVID-19 pandemic, it has become increasingly evident that a small but significant proportion of individuals experience persistent symptoms beyond the acute infection, a condition now widely referred to as long COVID or post-COVID syndrome (PCS). PCS is defined by the National Institute for Care and Excellence (NICE) as symptoms that exist three months after the acute infection^1^, and currently affects ∼2% of the UK population^2^.

Among the broad range of symptoms associated with PCS, fatigue is one of the most frequently reported and most disabling. In the United Kingdom, post-COVID fatigue (pCF) affects 71% of those with long COVID^2^, contributing to reduced occupational participation, diminished quality of life, and increased demand on healthcare services. The scale of this burden, combined with the heterogeneity of its presentation and the absence of specific diagnostic tools, underscores the urgency of understanding the biological underpinnings and clinical course of fatigue in this context.

Fatigue in PCS is multifactorial, encompassing not only overwhelming feelings of tiredness, but also cognitive/mental fatigue frequently described as “brain fog”^3^ and disproportionate symptom exacerbation after minimal exertion (post-exertional malaise)^4^. Reports also highlight muscle weakness, myalgia, and impaired motor coordination among long COVID patients^5–7^, indicating lasting muscle pathology even in individuals with a history of mild COVID-19^8–10^. The underlying mechanisms of pCF remain incompletely understood with hypotheses including immune dysregulation^11^, autonomic nervous system dysfunction^12^, mitochondrial impairment^10^, and psychosocial contributors^13^.

The severity and pattern of fatigue can vary widely between individuals, and its course may fluctuate over time. Some patients report gradual improvement or even complete remission^14^; others have persistent or relapsing fatigue over months to years.

To understand the clinical, biological and social dimensions of pCF, longitudinal studies are crucial. These allow for the identification of distinct trajectories and predictors of recovery or chronicity, by documenting clinical and physiological changes over time.

In a previous study^15^ comparing individuals with pCF to a healthy control cohort, we identified changes in physiology relating to dysautonomia, cortical excitability and muscle pathology. A subset of the same pCF cohort was then followed after 6 and 12 months^16^. As a cohort, most neurophysiological metrics as well as self-reported perception of fatigue significantly improved, slowly returning to levels seen in healthy controls. However, some individuals continue to suffer from post-COVID fatigue two or more years after their initial infection.

Here we take a larger cohort of individuals suffering from pCF after non-severe SARS-CoV-2 infection and demonstrate that neurophysiological measurements differ depending on the duration of pCF. We provide evidence that pCF is initially characterised by autonomic dysregulation, peripheral muscle pathology and reduced cortical excitability, in line with our previous results^15^. Over time, the neurophysiologic abnormalities evolve, with progressive changes in measures of cortical excitability and processing, as well as increasing peripheral muscle fatigue. By contrast, measures of autonomic function appear unchanged regardless of time since infection.

## Methods

### Participants

The study was approved by the Ethics Committee of Newcastle University Faculty of Medical Sciences. Participants provided written informed consent.

For this study, data from 145 participants (108 females) who were suffering from pCF by self-report were analysed. This included data from a pCF cohort of 37 participants (27 females) that were collected for our previous studies^15,16^ and baseline data (prior to any intervention) from individuals with pCF (108 participants, 81 females) that went on to participate in a vagus nerve stimulation trial (ISRCTN registry; ISRCTN18015802). For the electrophysiological tests, the previous cohort of 52 healthy volunteers (37 females) with no symptoms of fatigue served as the control dataset^15,16^. In this control cohort, six subjects had reported having mild COVID- 19 but with complete recovery and no symptoms of pCF.

All participants were 18-65 years of age and had no history of neurological disease.

For the purposes of this study, pCF participants were divided according to their time since infection (time of suffering from pCF), leading to two distinct groups: less than 200 days since COVID-19 infection (N=44; 31 females) and more than 200 days since infection (N=101; 77 females).

Figure 1 shows the time since infection for each individual in the pCF cohort.

**Figure 1:**
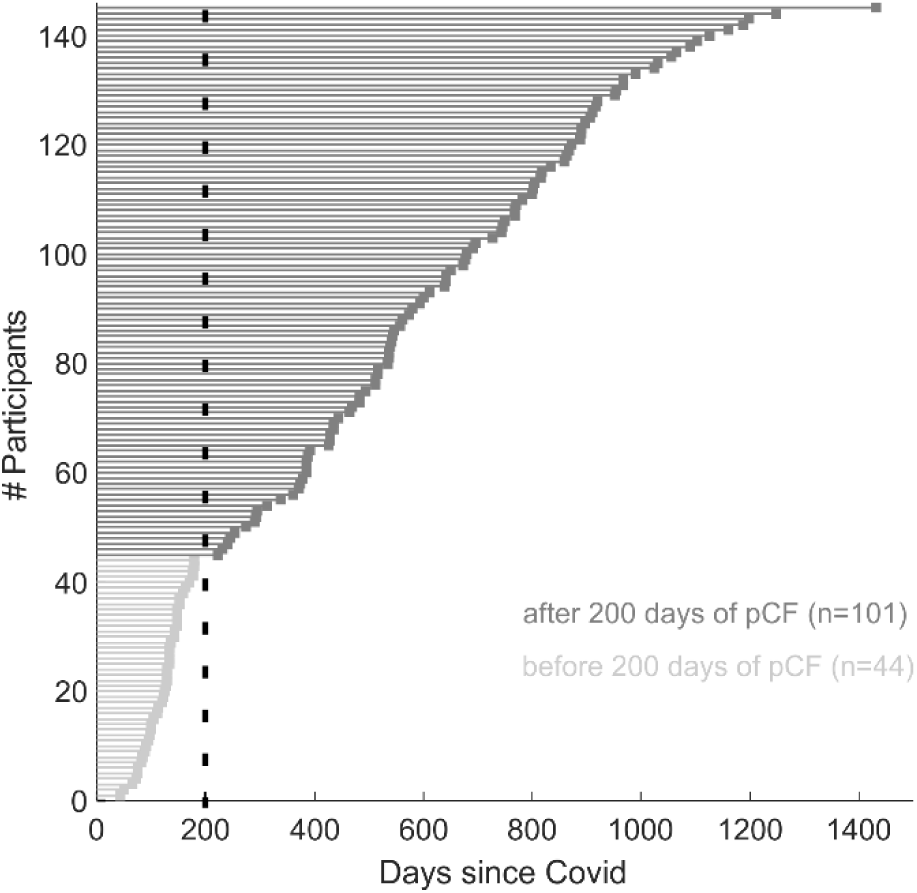
Distribution of time since SARS-CoV-2 infection which caused ongoing fatigue symptoms. *Light grey:* participants that had pCF for less than 200 days, *dark grey:* participants that had pCF for more than 200 days.

### Questionnaires

The Fatigue Impact Scale (FIS) was used to quantify subjective levels of fatigue. The FIS has 40 questions in total, evaluating the effect of fatigue on three domains of daily life: cognitive, physical and psychosocial functioning^17^.

A subset of 109 pCF participants also filled out a Fatigue Visual Analogue Scale, which simply asked them to score their fatigue that day on a scale from 0 (no fatigue) to 100 (extreme fatigue).

### General Electrophysiological Methods

The assessment protocols follow our previous studies^15,16^ which compared participants with pCF to age-matched controls and followed a cohort of pCF participants for a year. A summary of the various electrophysiological measures is illustrated in Figure 2.

**Figure 2:**
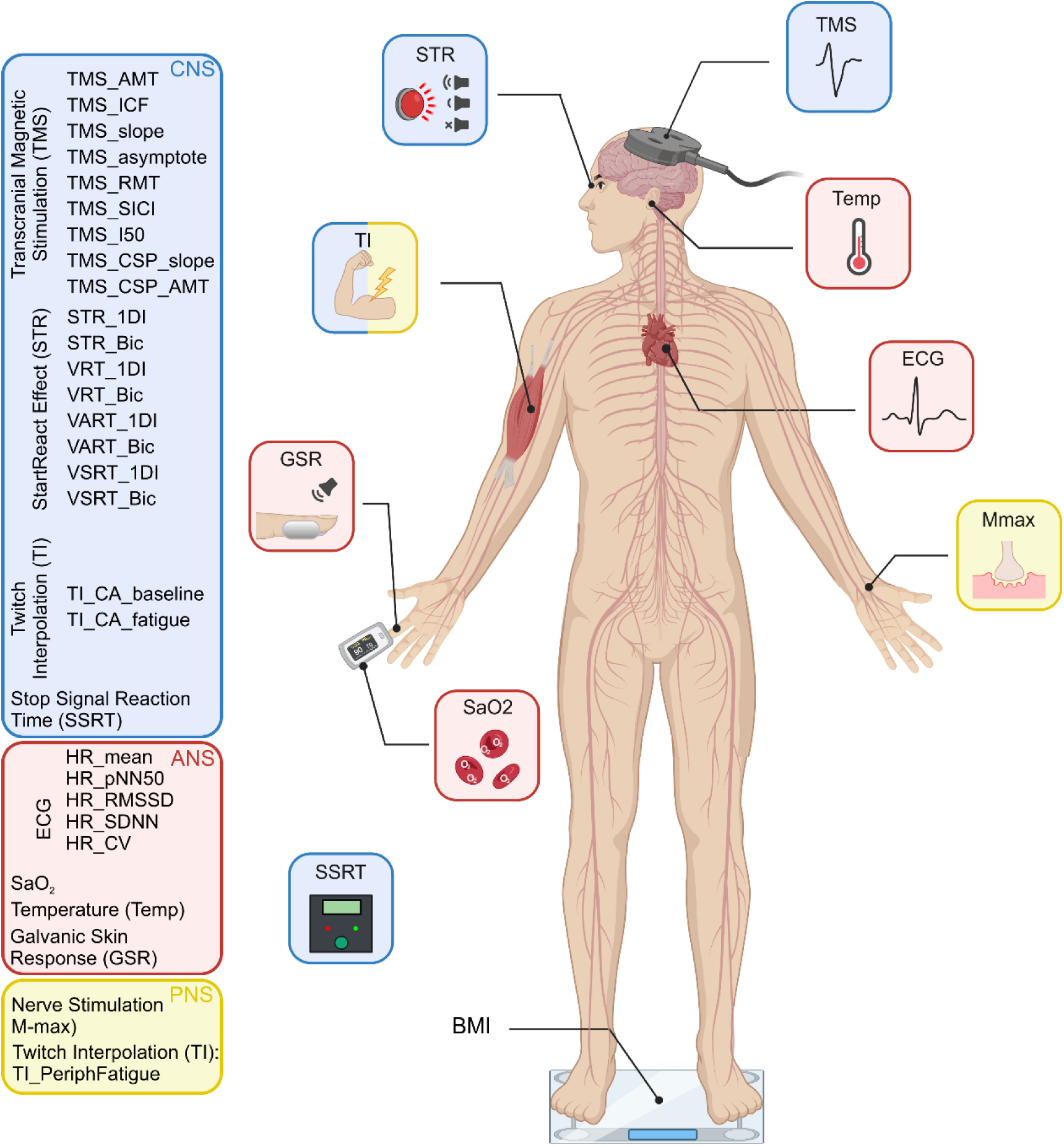
Schematic representation of the electrophysiological measures performed, separated according to which components of the nervous system (CNS, blue; PNS, yellow; and ANS, red) they assessed. Created with biorender.com.

Electromyogram (EMG) was recorded through adhesive surface electrodes placed on the skin over the muscle belly. EMG signals were amplified and filtered (band-width 30-2000 Hz) with a bioamplifier (D360 8-Channel Patient Amplifier, Digitimer, Welwyn Garden City, UK) and then converted to digital data with a sampling rate of 5kHz (CED Micro 1401 with Spike2 software, Cambridge Electronic Design, Cambridge, UK) and stored on a computer for off-line analysis. Where a measurement required a constant contraction, visual feedback of rectified and smoothed EMG activity was provided to the subject via a display of coloured bars on a computer screen, calibrated to the subject’s maximum voluntary contraction (MVC). Participants were then asked to maintain a contraction corresponding to 10% of their individual MVC. Transcranial magnetic stimuli were applied using a figure-of-eight coil through a Bistim 200^2^ stimulator (Magstim, Whitland, UK). The magnetic coil was held to induce electrical currents that flow perpendicular to the presumed line of the central sulcus in a posterior-anterior direction, with the handle pointing posteriorly and 45° away from the midline. The hotspot was defined as the region where the largest motor-evoked potential (MEP) in the target muscle could be evoked. To ensure a stable coil position during experiments and across sessions, the site of stimulation was marked using a Brainsight neuronavigation system (Rogue Research Inc., Montréal, Canada), which allows online navigation. A Polaris Vicra camera (Northern Digital Inc., Canada) was used to track the coil.

### Peripheral nerve stimulation

Stimuli to peripheral nerves (0.2 ms pulse width) were delivered using a Digitimer DS7AH isolated, constant current stimulator. The size of the maximal M wave was measured by stimulating the median nerve at the wrist and recording EMG from the abductor pollicis brevis (APB) muscle. Stimulus intensity was set to produce a supra-maximal M wave. Ten stimuli at this intensity were delivered and the highest amplitude M wave was used for subsequent normalisation of TMS recruitment curves (see below).

### TMS recruitment curve

As a measure of motor cortical excitability, the increase in APB MEP with increasing stimulus intensity was used^18^. The active motor threshold (AMT) was defined as the intensity which produces a MEP > 100 µV amplitude in at least 3 out of 6 stimuli, while the participant maintained an active contraction of 10% MVC. The intensity of the stimulation was expressed as a percentage of the maximal stimulator output (MSO). Recruitment curves of increasing intensities in 10% MSO steps were obtained in blocks of ten stimuli per step starting at AMT intensity, until 100% MSO was reached.

Offline analysis measured the size of MEPs from single trials and plotted this versus stimulus intensity. A sigmoid curve could then be fitted to the relationship^19^.

### Paired-pulse TMS

The hotspot was defined as the region where the largest MEP in the first-dorsal interosseous (1DI) muscle could be evoked with the minimum intensity. Resting motor threshold (RMT) was defined as the minimal stimulus intensity needed to produce a MEP > 100 µV amplitude in at least 3 out of 6 stimuli.

For the test stimulus, TMS intensity was adjusted to elicit MEPs of 1mV amplitude at rest, or to 120% RMT, whichever was lower. The conditioning stimulus intensity was set at 80% RMT.

Conditioning stimuli preceded the test stimuli by 3 or 10 ms and the recorded responses were compared to responses to the test stimulus alone, to measure short-interval intracortical inhibition (SICI) and intracortical facilitation (ICF) respectively Kujirai, et al. ^20^. Twenty stimuli for each condition were given in a pseudo-random order. Responses to the conditioning stimuli were expressed as a percentage to the responses to the test stimulus alone.

### Galvanic skin response

The galvanic skin response (GSR) is a measure of the cutaneous resistance or conductivity, which can be quantified by passing a weak electric current through a pair of electrodes. Variation in skin resistance depends on sweat production, which itself is mediated by the sympathetic system^21^; its habituation may be a relevant measure in assessing cognitive states^22^. The GSR was measured by placing two metal plates on the lateral and medial surfaces of the index finger. Five loud sounds (115 dB, C weighting, 500Hz, 50 ms, 8-8.8s inter-stimulus interval, chosen randomly from a uniform distribution) were played through loudspeakers placed underneath the subject chair, with the subject at rest. The ratio of the GSR response amplitude following the last stimulus compared to the first was used as a measure of habituation.

### StartReact effect

This paradigm measures reaction time from EMG in response to a visual cue (visual reaction time, VRT), a visual plus quiet auditory cue (visual-auditory reaction time, VART), and a visual plus loud auditory cue (visual-startle reaction time, VSRT). The acceleration of reaction time between VART and VSRT is termed the StartReact effect and has previously been used to assess connections from the reticulospinal system^23–26^.

A green light-emitting diode (LED) was located ∼1 m in front of the subject. Participants were instructed to flex their elbow and clench their fist as quickly as possible after the LED illuminated. EMG was recorded from both the first dorsal interosseous muscle (1DI) and biceps muscle, and reaction time measured as the time from cue to onset of the EMG burst. Three types of trial were randomly interleaved (20 repeats per condition; inter-trial interval 6-6.8s, interval chosen randomly from a uniform distribution): LED illumination alone (VRT), LED paired with a quiet sound (80dB, 500Hz, 50ms, VART), LED paired with a loud sound (500 Hz, 50ms, 115 dB, VSRT).

StartReact measurements were performed immediately after the GSR Habituation test, ensuring that any overt startle reflex had been habituated by the five loud sounds given in that test. The room lights were dimmed for this test.

Data were analysed offline trial-by-trial using a custom MATLAB program which identified the reaction time as the point where the rectified EMG exceeded the mean + 7 SD of the baseline measured 0-200 ms prior to the stimulus. Every trial was also inspected visually, and erroneous activity onset times (caused, for example, by electrical noise artefacts) was manually corrected. Average VRT, VART and VSRT were calculated for each subject and muscle, together with the amplitude of the StartReact effect, equal to VSRT-VART.

### Stop-signal reaction time

The stop-signal reaction time (SSRT) task measures the ability to inhibit a response after receiving a go cue. Participants are asked to respond to a go cue, but to inhibit their responses if a stop cue appears. This can indicate the state of premotor cortical-subcortical areas involved in impulse control^27^ and test for increased levels of inhibition in the motor system. SSRT was measured with a recently-developed portable device that uses Bayesian statistical analysis to improve the reliability of the measure^28^. The battery-powered device consisted of a plastic box which contained a microcontroller and a LCD screen, as well as one green LED, one red LED and a press button. Participants initiated a trial by pressing and holding down the response button. They were instructed to release this button as quickly as possible when the green LED was illuminated (GO trial; 75% of trials).

On 25% of trials, the red LED illuminated 5, 65, 135 or 195ms after the green LED (stop trial) and participants were instructed not to release the button on these trials. Trials were presented in three blocks of 64, with a 60 s break between each block. Each block consisted of 48 GO trials and 16 STOP trials (four for each delay). Using the distribution of reaction times on the GO trials, and the proportion of successfully inhibited responses, the algorithm calculated the SSRT as described in full in our previous work^28^.

### Electrocardiogram

A single channel of electrocardiogram (ECG) recording was captured, using a differential recording from left and right shoulder (bandpass 0.3-30 Hz) and stored for offline analysis. The time of each QRS complex was extracted. From these times, the mean heart rate and the pNN50 (a measure of heart rate variability and defined as the proportion of successive intervals which differ by >50ms) were computed. ECG was captured while participants were engaged in the SSRT test (see above), to ensure consistent behaviour across recordings.

### Twitch interpolation

Our previous studies^15,16^ used the twitch interpolation procedure, which allows assessment of an individual’s ability to activate a muscle voluntarily maximally, before and after a sustained (fatiguing) contraction. The experimental sequence followed previous work from this laboratory, to ensure that measurements would be comparable.

Subjects sat with their arm and forearm strapped into a dynamometer to measure torque about the elbow. The forearm was held vertically in supination, the upper arm horizontal and the elbow was flexed at a 90° angle. Velcro straps held the wrist, forearm and upper arm in place. Thin stainless-steel plate electrodes (30 x 15 mm) covered in saline-soaked gauze were used for electrical stimulation of the biceps muscle, by taping one electrode over the muscle belly and one over its distal tendon. The individual supramaximal stimulus level was set by increasing the intensity until the twitch response (recorded by the dynamometer) grew no further.

Upon receiving an auditory cue, subjects performed a 3s long maximal voluntary contraction until a stop tone sounded. Electrical stimulation to the biceps was given during MVC, 2s after the go cue and at rest, 5s after the stop cue. This sequence was repeated twice, with a 60 s rest period between go cues. After another 60s rest, another go cue then indicated to the subject to make a sustained MVC of up to 90s. If exerted force fell below 60% of the initial maximal level, contraction was terminated early. During this long MVC, the biceps muscle was stimulated every 10s. A final three stimuli (inter-stimulus interval 5s) were given at rest.

If a subject truly performs a maximal voluntary contraction, a superimposed electrical stimulus should not be capable of generating extra force. The size of any elicited twitch thus measures a central activation deficit. Stimulation of a fatigued muscle at rest after the long contraction produces a smaller twitch than that seen before the sustained MVC, indicating peripheral fatigue.

Central activation before and after fatigue, as well as peripheral fatigue were computed based on elicited twitches according to the calculations inMcDonald, et al. ^29^.

### Biometric Data

Various biometric measurements were collected from participants. These included blood oxygen saturation using a pulse oximeter placed onto the index finger, tympanic temperature, height, weight and full body composition (BC-545N Segmental Body Fat Scale, Tanita, Amsterdam, The Netherlands).

### Analysis

Data processing and statistical analyses were conducted using MATLAB (The MathWorks, Inc., Natick, MA, USA), SPSS (IBM SPSS Statistics for Windows, Armonk, NY: IBM Corp) and R (http://www.r-project.org/).

Descriptive statistics are given as mean ± standard deviation. Because each measure has different units and scales, data were normalized as a Z-score to allow easy comparison of differences. Z-scores were calculated by taking the difference in means of a measure between datasets (pCF vs healthy controls or pCF less than vs greater than 200 days) and dividing it by the standard deviation of the normative dataset (healthy controls or pCF less than 200 days). This is a measure of effect size and similar to Hedge’s g measure.

For the distribution of Z scores across subjects with similar times since infection, the sum of Z scores across all measures were taken for each subject. Subjects were ordered in ascending order according to their time since infection (most recently infected to longest time since infection) and groups of 20 subjects were taken to contribute to the overall sum of Z score for that bin, starting with the 20 most recently infected people. Each bin was moved by one subject at a time until the last bin included the 20 subjects that had pCF for the longest. If participants did not have a measure, the missing data was substituted by the mean of the Z scores across all subjects for that measure. For the sum of Z scores to reflect overall symptom burden, the signage of measures where the mean of the pCF cohort was smaller than the mean of the healthy control cohort was flipped (multiplied by -1), so that a higher Z score always reflected a larger distance from healthy for all measures.

To compare the cohorts (pCF vs healthy controls or the two separate pCF cohorts), unpaired t-tests were used. The Benjamini-Hochberg procedure was used to correct for multiple comparisons^30^. Raw (uncorrected) P-values are given throughout this report, but only stated as significant if they passed correction for multiple comparisons.

The correlation coefficient is given as Pearson’s r.

## Results

### Comparing Post-COVID Fatigue and Healthy Controls

Measures from all participants suffering from post-COVID fatigue (N=145; 108 female; 48.2 ± 9.8 years) were compared with a healthy control cohort (N=52; 37 females; 46.6 ± 9.4 years). Figure 3 illustrates results for all electrophysiological measures as a spider plot^31^, normalized as Z-scores and ordered so the greatest difference is located at the top of the figure. P-values are listed in Table 1.

**Figure 3:**
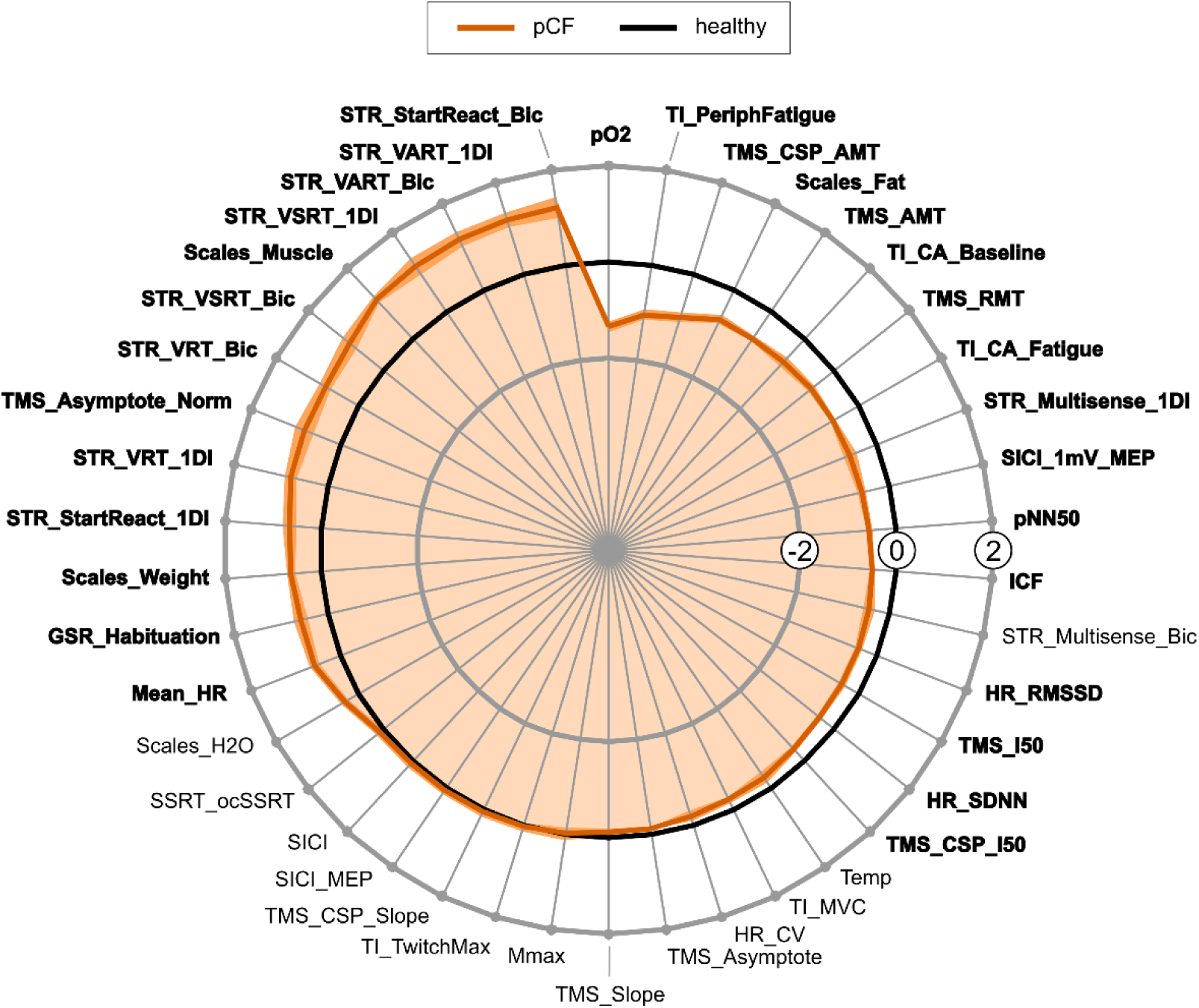
Comparison of pCF cohort with healthy controls. Data for each measure normalized as Z-scores (difference between pCF and control participants, scaled by SD). Black line at zero represents the control cohort, dark orange line the pCF cohort. Orange shading indicates the standard error of the mean difference (calculated by dividing the SD of each measure by the square root of the number of observations available). Measures are ordered so that the greatest difference is located at the top of the figure. Significant differences between the cohorts were assessed using unpaired t-tests. Measures passing the Benjamini-Hochberg correction for multiple comparisons are indicated in bold.

**Table 1:** List of p values for each measure comparing the whole pCF cohort (n=145) to the healthy control (n=52) cohort (left column), people who had pCF for less than 200 days (n=44) to healthy controls (middle left column), people who had pCF for over 200 days (n=101) to healthy controls (middle right column) and the two pCF cohorts to each other (right column). Measures between cohorts were compared with unpaired t-tests. Significant p-values that passed correction for multiple comparisons are marked in bold. P-values that were significant but did not pass correction for multiple comparisons are marked in grey. Unit of measure is given in brackets.

|  | All pCF vs healthy | < 200 days pCF vs healthy | > 200 days pCF vs healthy | Before vs after 200 days of pCF |
| --- | --- | --- | --- | --- |
| **FATIGUE IMPACT SCORE** |  |  |  |  |
| FIS_total | - | - | - | 0.002 |
| FIS_social | - | - | - | 0.003 |
| FIS_physical | - | - | - | 0.006 |
| FIS_cognitive | - | - | - | 0.010 |
| **CORTICAL** |  |  |  |  |
| SICI_RMT (%MSO) | 2.763x10<sup>-4</sup> | 0.536 | 1.071x10<sup>-6</sup> | 1.428x10<sup>-4</sup> |
| TMS_AMT (%MSO) | 5.887x10<sup>-7</sup> | 0.091 | 4.183x10<sup>-9</sup> | 6.328x10<sup>-5</sup> |
| TMS_I50 (%MSO) | 0.010 | 0.629 | 7.390x10<sup>-6</sup> | 3.451x10<sup>-6</sup> |
| TMS_asymptote (mV) | 0.130 | 0.407 | 0.204 | - |
| TMS_asymptote_norm (%Mmax) | 0.031 | 0.115 | 0.024 | 0.322 |
| TMS_slope (mV/%MSO) | 0.385 | 0.658 | 0.380 | - |
| TMS_CSP_AMT (ms) | 6.612x10<sup>-7</sup> | 0.057 | 4.117x10<sup>-9</sup> | 1.168x10<sup>-4</sup> |
| TMS_CSP_I50 (ms) | 0.013 | 0.388 | 3.935x10<sup>-6</sup> | 4.177x10<sup>-9</sup> |
| TMS_CSP_slope (ms/10%MSO) | 0.782 | 0.329 | 0.820 | - |
| SICI_ICF (%) | 4.795x10<sup>-5</sup> | 0.001 | 0.001 | 0.353 |
| SICI_SICI (%) | 0.725 | 0.755 | 0.748 | - |
| SICI_1mVMEP (%MSO) | 0.001 | 0.628 | 7.454x10<sup>-6</sup> | 4.225x10<sup>-4</sup> |
| SICI_MEP (mV) | 0.768 | 0.757 | 0.612 | - |
| TI_CA_baseline (%) | 0.002 | 0.230 | 0.001 | 0.018 |
| TI_CA_fatigued (%) | 4.005x10<sup>-6</sup> | 0.036 | 1.915x10<sup>-6</sup> | 0.006 |
| **REACTION TIME** |  |  |  |  |
| STR_VRT_Bic (ms) | 0.001 | 0.025 | 3.591 x10<sup>-4</sup> | 0.163 |
| STR_VRT_1DI (ms) | 4.604 x10<sup>-4</sup> | 0.017 | 2.622 x10<sup>-4</sup> | 0.252 |
| STR_VART_Bic (ms) | 3.056 x10<sup>-5</sup> | 0.022 | 3.405 x10<sup>-6</sup> | 0.008 |
| STR_VART_1DI (ms) | 7.676 x10<sup>-6</sup> | 0.011 | 7.392 x10<sup>-7</sup> | 0.010 |
| STR_VSRT_Bic (ms) | 0.003 | 0.274 | 0.001 | 0.015 |
| STR_VSRT_1DI (ms) | 4.778x10<sup>-4</sup> | 0.070 | 7.387 x10<sup>-5</sup> | 0.021 |
| STR_Multisens_Bic (ms) | 0.042 | 0.818 | 3.447 x10<sup>-4</sup> | - |
| STR_Multisens_1DI (ms) | 0.017 | 0.945 | 2.084 x10<sup>-5</sup> | 0.003 |
| STR_StartReact_Bic (ms) | 0.001 | 0.015 | 2.962 x10<sup>-4</sup> | 0.462 |
| STR_StartReact_1DI (ms) | 0.009 | 0.091 | 0.003 | 0.525 |
| SSRT_ocSSRT (ms) | 0.614 | 0.262 | 0.926 | - |
| **PERIPHERAL** |  |  |  |  |
| TI_PeriphFatigue (%) | 2.928 x10<sup>-7</sup> | 0.007 | 1.488 x10<sup>-8</sup> | 0.009 |
| TI_MVC (N) | 0.090 | 0.298 | 0.095 | - |
| TI_TwitchMax (N) | 0.838 | 0.035 | 0.492 | - |
| NMJ_Mmax (mV) | 0.919 | 0.359 | 0.809 | - |
| **AUTONOMIC** |  |  |  |  |
| Mean_HR (Hz) | 0.003 | 0.001 | 0.018 | 0.060 |
| HR_RMSSD (ms) | 0.007 | 0.007 | 0.027 | 0.347 |
| HR_SDNN (ms) | 0.004 | 0.015 | 0.012 | 0.488 |
| pNN50 (%ge) | 8.246 x10<sup>-6</sup> | 0.008 | 1.315 x10<sup>-5</sup> | 0.505 |
| HR_CV (Ratio) | 0.305 | 0.362 | 0.331 | - |
| pO2 (%) | 1.419 x10<sup>-10</sup> | 2.881 x10<sup>-7</sup> | 1.431 x10<sup>-9</sup> | 0.917 |
| GSR_Hab (Habituation %) | 0.022 | 0.068 | 0.021 | 0.786 |
| Temp (Co) | 0.272 | 0.057 | 0.044 | - |
| **BODY COMPOSITION** |  |  |  |  |
| Scales_Weight (kg) | 4.247 x10<sup>-4</sup> | 1.441 x10<sup>-4</sup> | 0.004 | 0.064 |
| Scales_Muscle (kg) | 3.416 x10<sup>-10</sup> | 3.770 x10<sup>-5</sup> | 1.959 x10<sup>-10</sup> | 0.306 |
| Scales_Fat (%) | 1.157 x10<sup>-4</sup> | 0.374 | 1.025 x10<sup>-6</sup> | 2.858 x10<sup>-4</sup> |
| Scales_H2O (%) | 0.045 | 0.984 | 0.006 | - |

Our results are largely consistent with our previously reported study comparing the same control cohort to a smaller subset of the pCF cohort used here^16^. For this present study we found several more metrics to be significantly different between patients with pCF and controls. This is most likely due to the increased number of subjects and the notably longer time this cohort had been suffering from the condition (505 ± 339 days since infection compared to 121 ± 37 days since infection for the previous cohort ^16^).

Multiple TMS measures related to cortical excitability were significantly different between the healthy control cohort and the pCF cohort (resting motor threshold *TMS_RMT* p<0.001, active motor threshold *TMS_AMT* p<0.001, the asymptote of the recruitment curve normalized to M-max *TMS_Asymptote_Norm* p=0.031, the intensity yielding 50% of the asymptote response amplitude *TMS_I50* p=0.0103, active motor threshold of the cortical silent period *TMS_CSP_AMT* p<0.001 and I50 of the cortical silent period *TMS_CSP_I50* p=0.0133).

The recruitment curve slope (*TMS_Slope* p=0.385), the asymptote of the recruitment curve (*TMS_Asymptote* p=0.130) and the recruitment curve slope of the cortical silent period (*TMS_CSP_Slope* p=0.782) were not significantly different between cohorts. Additionally, intracortical facilitation was significantly decreased in pCF patients (*ICF* p<0.001), but short-interval intracortical inhibition did not show a difference between patients with pCF and controls (*SICI* p=0.725).

We found longer reaction times in pCF versus controls for all measures relating to the StartReact paradigm in both muscles (visual reaction time *STR_VRT_Bic* p<0.001 and *STR_VRT_1DI* p<0.001, visual-auditory reaction time *STR_VART_Bic* p<0001 and *STR_VART_1DI* p<0.001, visual-startle reaction time *STR_VSRT_Bic* p<0.003 and *STR_VSRT_1DI* p<0.001). The StartReact effect, which measures the acceleration of a visual reaction time by a loud (startling) sound and has been proposed to assess reticulospinal pathways^32^, was also significantly increased in both muscles (*STR_StartReact_Bic* p<0.001 and *STR_StartReact_1DI* p=0.009), while the difference between visual reaction time and visual-auditory reaction time was reduced in pCF versus controls (*STR_Multisense_1DI* p=0.0171) but did not pass correction for multiple comparisons in the biceps muscle (*STR_Multisense_Bic* p=0.042). The stop-signal-reaction time (*SSRT_ocSSRT* p=0.614) was the same for both cohorts.

We also found evidence of autonomic dysregulation in the pCF group, showing lower peripheral blood oxygen saturation (*pO2* p<0.001), lower galvanic skin response habituation (*GSR_Habituation* p=0.022), increased resting heart rate (*Mean_HR* p=0.003) and lower heart rate variability (*pNN50* p<0.001, *HR_RMSSD* p=0.007, *HR_SDNN* p=0.004). Other measures of autonomic function (tympanic temperature *Temp* p=0.272 and variability in heart rate coefficient of variation *HR_CV* p=0.305) did not differ between cohorts.

Maximal voluntary contraction (*TI_MVC* p=0.090) was not significantly reduced in pCF, suggesting no deficit in force production for brief contractions. There also was no difference in maximal M-wave (*Mmax* p=0.919), or their maximal elicited muscle twitch (*TI_TwitchMax* p=0.838). However, after a prolonged maximal contraction, pCF participants had an increased level of peripheral fatigue (size of maximum twitch evoked by direct electrical stimulation of the muscle after a sustained contraction compared with baseline; *TI_PeriphFatigue* p<0.001).

Central activation, which assesses the ability of the CNS to activate muscle maximally voluntarily, was significantly reduced in pCF, either assessed at baseline (*TI_CA_baseline* p=0.002) or after a fatiguing contraction (*TI_CA_fatigue* p<0.001).

In terms of body composition, pCF participants weighed significantly more (*Scales_Weight* p<0.001), but their body weight actually consisted of more muscle mass (*Scales_Muscle* p<0.001) and less fat by percentage (*Scales_Fat* p<0.001), whereas there was no difference in water mass by percentage (*Scales_H2O* p=0.045, not significant after correcting for multiple comparison) between the cohorts.

### Changes in Physiology over time

#### Sum of Z Scores Distribution

To assess physiological differences over time, a sum of Z scores across all measures was calculated for each subject. Subjects were ordered in ascending order according to their time since infection (most recently infected to longest time since infection) and bins of 20 subjects were taken to contribute to the overall sum of Z score for that bin, starting with the 20 most recently infected people. Each bin was moved by one subject at a time until the last bin included the 20 subjects that had pCF for the longest. Figure 4A illustrates sum of Z scores for these bins over time, with time since COVID for each bin being the average time since infection across the 20 people in that bin.

**Figure 4:**
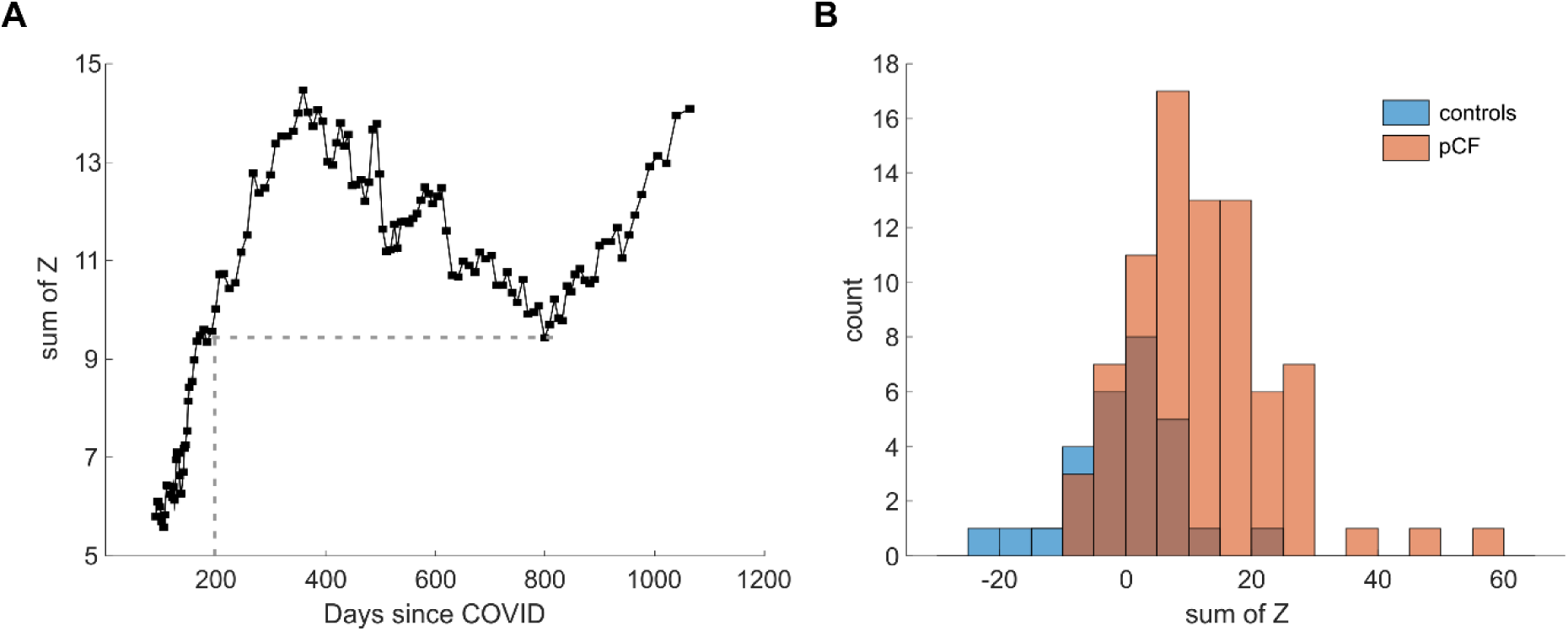
Distribution of sum of Z scores across all measures. **A:** Each data point represents a bin of 20 subjects. Subjects were ranked in ascending order according to their time since infection (most recently infected to longest time since infection) and bins of 20 subjects were taken to contribute to the overall sum of Z score for that bin, starting with the 20 most recently infected people. Each bin was moved by one subject at a time until the last bin included the 20 subjects that had pCF for the longest. If participants did not have a measure, the missing data were substituted by the mean of the Z scores across all subjects for that measure. For the sum of Z scores to reflect overall symptom burden, the signage of measures where the mean of the pCF cohort was smaller than the mean of the healthy control cohort was flipped (multiplied by -1), so that a higher Z score always reflected a larger distance from healthy controls for all measures. **B:** Sum of Z scores for each subject of the pCF (orange) and healthy control (blue) cohorts. The histogram has a bin width of 5.

The sum of Z rises steeply between 90 and 360 days since infection, indicating an increase in physiological differences compared to healthy controls over that time period. From 360 days to 800 days since infection, the sum of Z then drops about halfway before rising again to similar levels as the peak at 360 days. The midpoint of the rising curve (grey dotted lines in Figure 4A) was determined as 200 days since COVID infection. We chose this cut off point, because all bins with more than 200 days since COVID have higher sum of Z than those with less than 200 days since COVID.

Figure 4B validates the sum of Z scores as a measure of overall symptom burden by illustrating its distribution among the healthy control cohort and the pCF cohort. While there is some overlap between the two cohorts, the pCF cohort shows a clear shift towards higher sums of Z scores across measures (p<0.001).

#### Differences in Individuals Before and After 200 Days of Post-COVID Fatigue

Participants were split into two groups according to their time since infection. The first group contained people that had pCF for less than 200 days (N=44; 31 females; 46.7 ± 9.1 years) and the second group contained people that had pCF for over 200 days (N=101; 77 females; 48.8 ± 10.1 years). We refer to these groups below using the shorthand pCF<200 and pCF>200.

Measures that were significantly different between pCF and the control cohort (Figure 3) were compared with unpaired t-tests between pCF<200 and pCF>200. Figure 5 illustrates results for these electrophysiological measures as a spider plot^31^, normalized as Z-scores and ordered so the greatest difference is located at the top of the figure. P-values are listed in Table 1.

**Figure 5:**
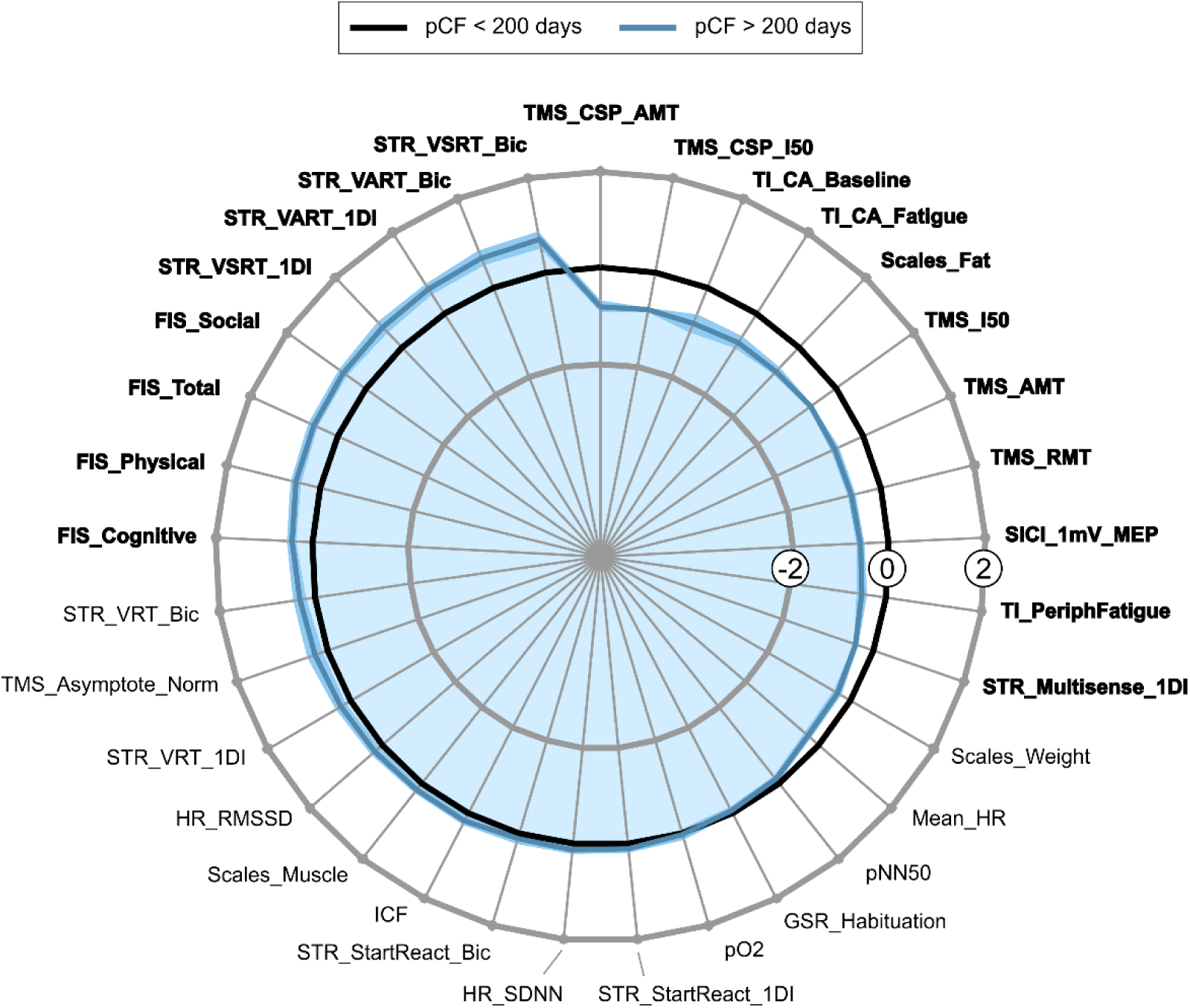
Comparison of participants that had pCF for less than 200 days (black) and over 200 days (blue). Data for each measure normalized as Z-scores. Black line at zero represents the pCF cohort whose infection was less than 200 days ago, dark blue line the pCF cohort who was infected over 200 days ago. Blue shading indicates the standard error of the mean difference (calculated by dividing the SD of each measure by the square root of the number of observations available). Measures are ordered so that the greatest difference is located at the top of the figure. Significant differences between the cohorts were assessed using unpaired t-tests. Measures passing the Benjamini-Hochberg correction for multiple comparisons are indicated in bold.

pCF>200 participants reported significantly higher levels of fatigue according to the Fatigue Impact Scale compared pCF<200 (*FIS_Total* pCF<200 85.5 ± 25.8, pCF>200 100.0 ± 25.4, p=0.002). pCF>200 scored notably higher on all FIS subscores (*FIS_Social* pCF<200 38.8 ± 13.7, pCF>200 46.6 ± 14.3, p=0.003; *FIS_Physical* pCF<200 24.8 ± 5.6, pCF>200 27.8 ± 6.0, p=0.006; *FIS_Cognitive* pCF<200 21.9 ± 8.7, pCF>200 25.6 ± 7.6, p=0.011).

pCF>200 also had higher peripheral muscle fatigue (*TI_PeriphFatigue* p=0.009) and a lower body fat percentage (*Scales_Fat* p<0.001).

Importantly, all other measures that were significantly different between pCF<200 and pCF>200 assessed the central nervous system. These included TMS measures showing increased cortical excitability in pCF>200 (*TMS_AMT* p<0.001; *TMS_RMT* p<0.001; *TMS_I50* p<0.001; *TMS_CSP_AMT* p<0.001; *TMS_CSP_I50* p<0.001; *SICI_1mV_MEP* p<0.001), significantly slower reaction times in pCF>200 (*STR_VART_1DI* p=0.010; *STR_VSRT_Bic* p=0.015; *STR_VSRT_1DI* p=0.021; *STR_Multisens_1DI* p=0.003) and a larger central activation deficit in pCF>200 (*TI_CA_Fatigue* p=0.006; *TI_CA_Baseline* p=0.018).

Correlating neurophysiological measures with time since infection (Supplementary Table 1), we found significant correlations mainly for measures of cortical excitability (*TMS_AMT (p<0.001), TMS_RMT (p=0.001), TMS_1mV_MEP (p=0.002), TMS_I50 (p<0.001), TMS_asymptote (p<0.001), TMS_CSP_AMT (p<0.001)* and *TMS_CSP_I50 (p<0.001)*, as well as for the FIS scores (*FIS_total, FIS_social, FIS_cognitive, FIS_physical*), measures of peripheral muscle function (*TI_PeriphFatigue, TI_TwitchMax*), blood oxygen saturation (*pO_2_*) and body temperature (*temp*); all p<0.001).

However, measures assessing autonomic function of the nervous system seemed to be similar between pCF<200 and pCF>200 (*Mean_HR* p=0.060; *HR_RMSSD* p=0.347; *HR_SDNN* p=0.489; *pNN50* p=0.505; *GSR_Habituation* p=0.786; *pO_2_* p=0.918). There was also no difference in intracortical facilitation (*ICF* p=0.353), or in body weight and muscle percentage (*Scales_Weight* p=0.064; *Scales_Muscle* p=0.306) and some measures of reaction time such as the StartReact effect were also similar (*STR_StartReact_Bic* p=0.462; *STR_StartReact_1DI* p=0.525; *STR_VRT_Bic* p=0.163; *STR_VRT_1DI* p=0.252).

More evidence that people suffering from pCF for over 200 days deteriorate physiologically comes from comparing each pCF cohort to healthy controls separately (Figure 6). Figure 6A compares pCF<200 with healthy controls; only 8 measures out of 42 showed a significant difference (P-values are listed in Table 1). These measures included those which we previously reported to be different between healthy controls and pCF^16^, including intracortical facilitation (*ICF* p=0.001), mean resting heart rate (*Mean_HR* p=0.015), peripheral muscle fatigue (*TI_PeriphFatigue* p=0.007) and blood oxygen (*pO_2_* p<0.001). Additionally, here we also found reduced heart rate variability (*HR_RMSSD* p=0.007; *pNN50* p=0.008) and a difference in body composition (*Scales_Muscle* p<0.001; *Scales_Weight* p<0.001).

**Figure 6:**
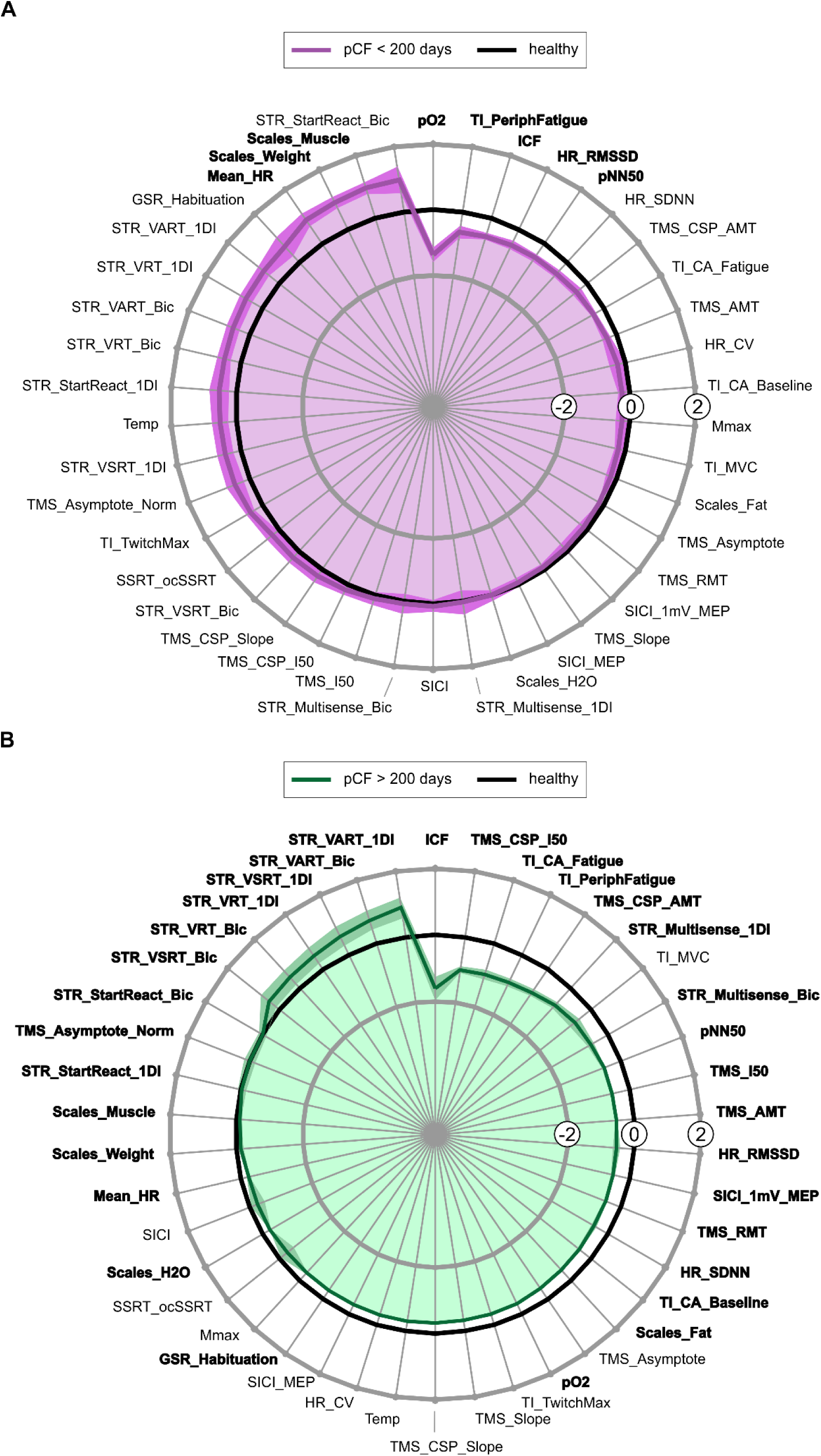
Comparison of the two pCF cohorts with healthy controls. Data for each measure are normalized as Z-scores. Measures are ordered so that the greatest difference is located at the top of the figure. Significant differences between the cohorts were assessed using unpaired t-tests. Measures passing the Benjamini-Hochberg correction for multiple comparisons are indicated in bold. **A:** Black line at zero represents the control cohort, dark purple line the pCF cohort who was infected less than 200 days ago. Purple shading indicates the standard error of the mean difference. **B:** Black line at zero represents the control cohort, dark green line the pCF cohort for whom the index infection occurred over 200 days before recruitment. Green shading indicates the standard error of the mean difference.

When comparing pCF>200 with healthy controls (Figure 6B), it became evident that they deviated more from healthy controls, with 31 out of 42 measures being significantly different. P-values are listed in Table 1.

In pCF>200, two more measures relating to autonomic function were significantly different from healthy controls (*GSR_Habituation* p=0.021; *HR_SDNN* p=0.012) in addition to the four measures of the group with pCF for a shorter time (*pO_2_* p<0.001; *Mean_HR* p=0.018; *HR_RMSSD* p=0.027; *pNN50* p<0.001). This pCF cohort also showed a central activation deficit (*TI_CA_Fatigue* p<0.001; *TI_CA_Baseline* p<0.001) as well as the peripheral muscle fatigue (*TI_PeriphFatigue* p<0.001) already present in the more recently infected cohort. There were more changes in body composition (*Scales_Fat* p<0.001; *Scales_H_2_0* p=0.006; as well as *Scales_Muscle* p<0.001; *Scales_Weight* p=0.004). In addition to increased cortical facilitation (*ICF* p<0.001) this cohort showed changes in a multitude of measures assessing cortical excitability (*TMS_CSP_AMT* p<0.001; *TMS_AMT* p<0.001; *TMS_RMT* p<0.001; *TMS_CSP_I50* p<0.001; *TMS_I50* p<0.001; *SICI_1mV_MEP* p<0.001; *TMS_asymptote_norm* p=0.024). Lastly, people that had pCF for over 200 days also had decreased reaction times compared to healthy controls (*STR_VART_1DI* p<0.001; *STR_VART_Bic* p<0.001; *STR_Multisens_1DI* p<0.001; *STR_VSRT_1DI* p<0.001; *STR_VRT_1DI* p<0.001; *STR_Startreact_Bic* p<0.001; *STR_Multisens_Bic* p<0.001; *STR_VRT_Bic* p<0.001; *STR_VSRT_Bic* p<0.001; *STR_StartReact_1DI* p=0.003), which was not seen in people that had pCF for less than 200 days.

## Discussion

Long COVID has emerged as a debilitating and common complication of the COVID-19 pandemic, which can affect people even after a mild initial infection. Studies indicate that approximately 10-20% of people infected with SARS-CoV-2 continue to experience long-term sequelae up to six months after the acute infection^33,34^. In the UK, an estimated 3.1% of the population were experiencing persistent symptoms after a COVID infection^2^. While some individuals affected by post-COVID syndrome will spontaneously recover, there is currently no clear treatment for the condition and many people continue to suffer from PCS years after their infection. Moreover, it is far from clear why some individuals recover spontaneously over time, while others show no improvement, or worse, deteriorate.

Access to this large dataset has allowed us to explore neurophysiological differences between individuals with post-COVID fatigue, according to the length of time individuals have been affected by the condition. Within the first 6 months of having pCF, individuals already show a number of changes in all three main divisions of the nervous system, in line with our previous results^15^. People whose symptoms persist for longer than 6 months report significantly higher levels of fatigue; this is corroborated by objective changes in peripheral muscle fatigue and measures of cortical excitability and processing.

To assess overall physiological changes over time, the sum of Z scores across all measures was taken. Individuals with pCF tended to have a higher sum than healthy controls (p<0.001; Figure 4B) and there was a sharp incline between people having had pCF for three months (definition of PCS according to NICE guidelines) to approximately one year (Figure 4A). Based on this, we divided the pCF cohort into two distinct groups, in order to investigate which measures were driving this worsening (diverging further away from healthy controls) over time.

The first group contains people that have been affected by pCF for less than 200 days. When comparing this cohort to healthy controls, we found evidence for autonomic dysfunction (decreased blood oxygen saturation, increased heart rate and decreased heart rate variability), increased peripheral muscle fatigue (*TI_PeriphFatigue*) and a reduction in intra- cortical facilitation (*ICF*), a measure of intracortical glutamatergic function. There was also an increase in body weight, though notably it was mainly the percentage of muscle mass that appeared to increase.

These results largely replicate our previous findings^15^, which included individuals that had pCF for up to 179 days back in 2021. The addition of people that had pCF for less than 200 days, but caught COVID roughly two years later in 2023, did not change these initial results, which suggests that different virus variants have largely the same effect on pCF. The cohort of individuals that had pCF for over 200 days at the time of testing includes people that were infected in late 2020 and up to early 2023. From alpha to omicron, the UK would have had 6 different virus variants during that time^35^, with several subvariants. It is impossible to say for certain who caught which specific variant, therefore we cannot draw any conclusions on potential variant-specific effects.

People who continue to suffer from pCF after 200 days might reflect a group of true “long-haulers”, with a spontaneous recovery becoming less likely after that time. These individuals report a significant increase in fatigue (FIS scores) compared to people that had pCF for a shorter amount of time. Objective measures support the fact that these individuals are left worse off over time, with several measures of cortical excitability and cognitive processing as well as peripheral muscle fatigue significantly changed after 200 days of pCF.

Neuromuscular fatigue is defined as a reduction in the capacity of muscles to generate force and has both peripheral and central origins. Peripheral fatigue reflects a failure of the peripheral nervous system, whereas central fatigue refers to the reduced ability of the central nervous system to activate muscles fully. During a sustained, fatiguing contraction, cortical excitatory drive increases in order to maintain force output, but inhibitory mechanisms such as intracortical inhibition and CSP concurrently intensify, resulting in an overall diminished corticospinal efficacy^36^.

After 200 days of being affected by pCF, people show increased cortical excitability, with lower motor thresholds (*TMS_RMT, TMS_AMT, TMS_CSP_AMT*) and less stimulator intensity required to achieve 50% of both maximal MEP amplitude and maximal CSP duration (*TMS_I50, TMS_CSP_I50*), compared to individuals that had pCF for less than 200 days (Figure 5). Despite this cortical over-excitability, the cohort also had impaired central activation (*TI_CA_baseline, TI_CA_Fatigue*), indicating a deficit in the effectiveness of their cortical output.

This elevation of corticomotor excitability is likely a compensatory adjustment of the central motor networks, attempting to preserve force output despite reduced peripheral capacity to generate force. Peripheral myopathic changes are already apparent in individuals that had pCF for less than 200 days, whose muscles show a reduced ability to produce force in response to electrical stimulation after a sustained contraction (*TI_PeriphFatigue*). This muscle fatigue is further increased after 200 days of having pCF.

Emerging evidence suggests that mitochondrial dysfunction is a potential mechanism for these changes. SARS-CoV-2 may impair mitochondrial function in muscles directly^37^, even in individuals suffering from PCS that have not been hospitalised^9,10^. Additionally, SARS-CoV-2 infection alters myokine signalling to sustain inflammation^38,39^, which also impairs mitochondrial function^40^.

Individuals that had pCF for longer than 200 days also had significantly slower reaction times (*STR_VART_Bic, STR_VART_1DI, STR_VSRT_Bic, STR_VSRT_1DI*), indicating diminished cognitive processing^41^, in line with increased cognitive fatigue.

Notably, none of the measures of autonomic function were different between the two pCF cohorts. Autonomic dysregulation is a well-established feature of chronic fatigue^42^ and multiple studies indicate autonomic dysfunction after COVID-19^12,43–46^. Supporting this, we found multiple abnormalities in autonomic function when comparing individuals with pCF to healthy controls. Even before 200 days, people with pCF have reduced blood oxygen saturation (*pO_2_*), increased resting heart rate (*mean_HR*) and reduced heart rate variability (*HR_RMSSD, pNN50*). While people affected by pCF for longer than 200 days also show reduced blood oxygen saturation and heart rate variability (*HR_RMSSD, pNN50, HR_SDNN*), surprisingly, their resting heart rate is lower than healthy controls, and habituation of their galvanic skin response (GSR_Habituation) is increased (Figure 6B).

This suggests that while some dysautonomia is present both before and after 200 days of pCF, it appears to remain stable, unlike measures of cortical excitability and cognitive processing, which worsen over time. The changes in resting heart rate and galvanic skin response habituation could reflect either a normalisation of underlying physiological function, or a secondary compensatory process in which the primary pathology is masked by homeostatic adjustments.

When looking at the distribution of the sum of Z scores over time, there is a drop between 360 and 800 days since infection, before it rises again to similar levels as the peak at 360 days (Figure 4A). This pattern of rising and falling could imply a general fluctuation in symptom burden, or perhaps this is a reflection of ongoing physiological adaptations. Longer follow up studies are needed to determine how these trends continue after 1200 days.

In our previous longitudinal study^16^, we followed 18 participants with pCF over 12 months and found most neurophysiological metrics tended to return slowly to levels seen in healthy controls, as well as participants reporting decreasing levels of fatigue. The progressive normalization of aberrant measures, in parallel with improvements in Fatigue Impact Scores, suggests that these changes arise as a consequence of a SARS-CoV-2 infection, rather than reflecting a pre-existing phenotype predisposing individuals to develop post-COVID fatigue^16^.

These observations, together with our findings here that people suffering from pCF for longer show a general worsening of measures, indicates that while there is improvement over time on an individual level, those people who remain fatigued beyond a critical time point develop mal-adaptations (or perhaps fail to adapt) that contribute to them feeling physically, cognitively and socially more fatigued than when the symptoms of post-COVID fatigue initially emerged. A recent study by Taquet et al.^47^ found very similar cognitive and psychiatric symptom trajectories over 3 years. Their analysis of questionnaire scores concludes that symptoms at 2-3 years were strongly predicted by the degree of recovery at 6 months, with people with severe scores at 6 months very likely to experience an increase in symptom burden over time. This further corroborates 6 months (∼ 200 days) as the critical time point beyond which accumulating physiological changes start to exacerbate PCS symptoms.

Our findings have implications for other studies aiming to investigate the mechanisms behind post-COVID fatigue and the development of treatment approaches. Although we cannot say what effects different variants or predispositions might have on the condition, it is evident that people deviate physiologically based on how long they have been afflicted by pCF. This could complicate comparative interpretation of different studies, since a pCF cohort meeting the definition for pCF (3 months of persistent symptoms) potentially deviates significantly from individuals with ongoing pCF over years. A key challenge for future trials will be to balance recruitment to ensure even representation of pCF phenotypes. Without deliberate stratification of symptom duration or physiological profile, treatment effects risk being diluted or misinterpreted.

## Data Availability

All data produced in the present study are available upon reasonable request to the authors

