## Supplementary Table 1 for "Evidence of Accumulating Neurophysiologic Dysfunction in Persistent Post-COVID Fatigue"

### Supplementary Information

| **measure** | **p** | **r** |
|---|---|---|
| pO2 (%) | **1.33E-05** | **0.3531** |
| TMS_CSP_I50 (ms) | **3.88E-05** | **-0.3347** |
| Temp (Co) | **2.05E-04** | **-0.3036** |
| TMS_CSP_AMT (ms) | **8.58E-04** | **-0.2739** |
| FIS_total | **0.0015** | **0.2615** |
| TMS_AMT (%MSO) | **0.0024** | **-0.2507** |
| FIS_social | **0.0031** | **0.2441** |
| FIS_physical | **0.0032** | **0.2429** |
| TMS_I50 (%MSO) | **0.0034** | **-0.2418** |
| TMS_asymptote (mV) | **0.0042** | **-0.2362** |
| TI_PeriphFatigue (%) | **0.0043** | **-0.2357** |
| FIS_cognitive | **0.0053** | **0.2305** |
| TI_TwitchMax (N) | **0.0067** | **-0.2242** |
| SICI_RMT (%MSO) | **0.0112** | **-0.2102** |
| SICI_1mVMEP (%MSO) | **0.0147** | **-0.2023** |
| STR_StartReact_Bic (ms) | 0.0468 | 0.1654 |
| STR_VART_Bic (ms) | 0.0515 | 0.1621 |
| STR_VART_1DI (ms) | 0.0588 | 0.1573 |
| STR_Multisens_Bic (ms) | 0.0736 | -0.1490 |
| TMS_CSP_slope (ms/10%MSO) | 0.0770 | -0.1473 |
| TI_CA_fatigued (%) | 0.0854 | -0.1434 |
| STR_Multisens_1DI (ms) | 0.0882 | -0.1421 |
| HR_CV (Ratio) | 0.0924 | -0.1403 |
| TMS_asymptote_norm (%Mmax) | 0.1152 | -0.1314 |
| pNN50 (%ge) | 0.1472 | -0.1210 |
| GSR_Hab (Habituation %) | 0.1605 | -0.1171 |
| STR_VSRT_1DI (ms) | 0.1821 | 0.1114 |
| STR_StartReact_1DI (ms) | 0.1986 | 0.1074 |
| STR_VRT_Bic (ms) | 0.2342 | 0.0994 |
| STR_VSRT_Bic (ms) | 0.2645 | 0.0933 |
| STR_VRT_1DI (ms) | 0.2714 | 0.0919 |
| TMS_slope (mV/%MSO) | 0.2989 | -0.0869 |
| SICI_MEP (mV) | 0.4099 | 0.0690 |
| SSRT_ocSSRT (ms) | 0.4281 | 0.0663 |
| NMJ_Mmax (mV) | 0.4838 | 0.0586 |
| TI_CA_baseline (%) | 0.5812 | -0.0462 |
| Scales_Weight (kg) | 0.6419 | -0.0389 |
| TI_MVC (N) | 0.6829 | -0.0342 |
| HR_SDNN (ms) | 0.7169 | -0.0304 |
| Scales_Muscle (kg) | 0.7524 | 0.0264 |
| HR_RMSSD (ms) | 0.8062 | -0.0206 |
| SICI_ICF (%) | 0.8598 | 0.0148 |
| Mean_HR (Hz) | 0.8667 | -0.0141 |
| Scales_H2O (%) | 0.9294 | 0.0074 |
| Scales_Fat (%) | 0.9499 | -0.0053 |
| SICI_SICI (%) | 0.9596 | 0.0042 |

**Supplementary Table 1:** Correlations between neurophysiological measures and time since infection. Pearson's correlation coefficient *r* and associated p values are reported, in ascending order from smallest to largest p value. Significant correlations ($p < 0.005$) that pass Benjamini-Hochberg correction for multiple comparisons are highlighted in bold. Unit of measure is given in brackets.
